# Renal Outcomes of Staged Versus Concomitant Percutaneous Coronary Intervention and Transcatheter Aortic Valve Replacement: A Systematic Review and Meta-Analysis

**DOI:** 10.64898/2026.07.19.26353414

**Authors:** Veda Chanda, Vinicius Bittar, Pedro E. P. Carvalho, Philippe Garot

## Abstract

**Background:** The optimal timing of percutaneous coronary intervention (PCI) in patients undergoing transcatheter aortic valve replacement (TAVR) remains unclear, particularly regarding its impact on renal outcomes.

**Methods:** We conducted systematic review and meta-analysis of studies comparing staged versus concomitant PCI in patients with aortic stenosis and coronary artery disease undergoing TAVR. We searched MEDLINE, Embase, and Cochrane databases comprehensively. Using a random-effects model, we calculated odds ratios (OR) with 95% confidence intervals (CI) to assess the incidence of contrast-induced acute coronary injury (CI-AKI) across different stages.

**Results:** The analysis included 11 studies encompassing 7,119 patients. Overall, staged PCI did not significantly differ from concomitant PCI in reducing CI-AKI (OR 1.02; 95% CI 0.53 to 1.98; p = 0.959; Figure 1A). Subgroup analysis revealed no significant differences in stage 1 (OR 1.99; 95% CI 0.38 to 10.47; p = 0.417; Figure 1B) or stage 2 CI-AKI (OR 1.01; 95% CI 0.39 to 2.64; p = 0.978; Figure 1C). However, a statistically significant difference emerged for stage 3/4 CI-AKI, favoring the staged approach (OR 0.48; 95% CI 0.24 to 0.99; p = 0.046; Figure 1D).

Figure 1.
Primary Endpoints for CI-AKI **Legend:** Compared to concomitant procedures, PCI and TAVI in a staged approach significantly reduced the odds of stages 3 or 4 CI-AKI in patients with CAD and SAS.

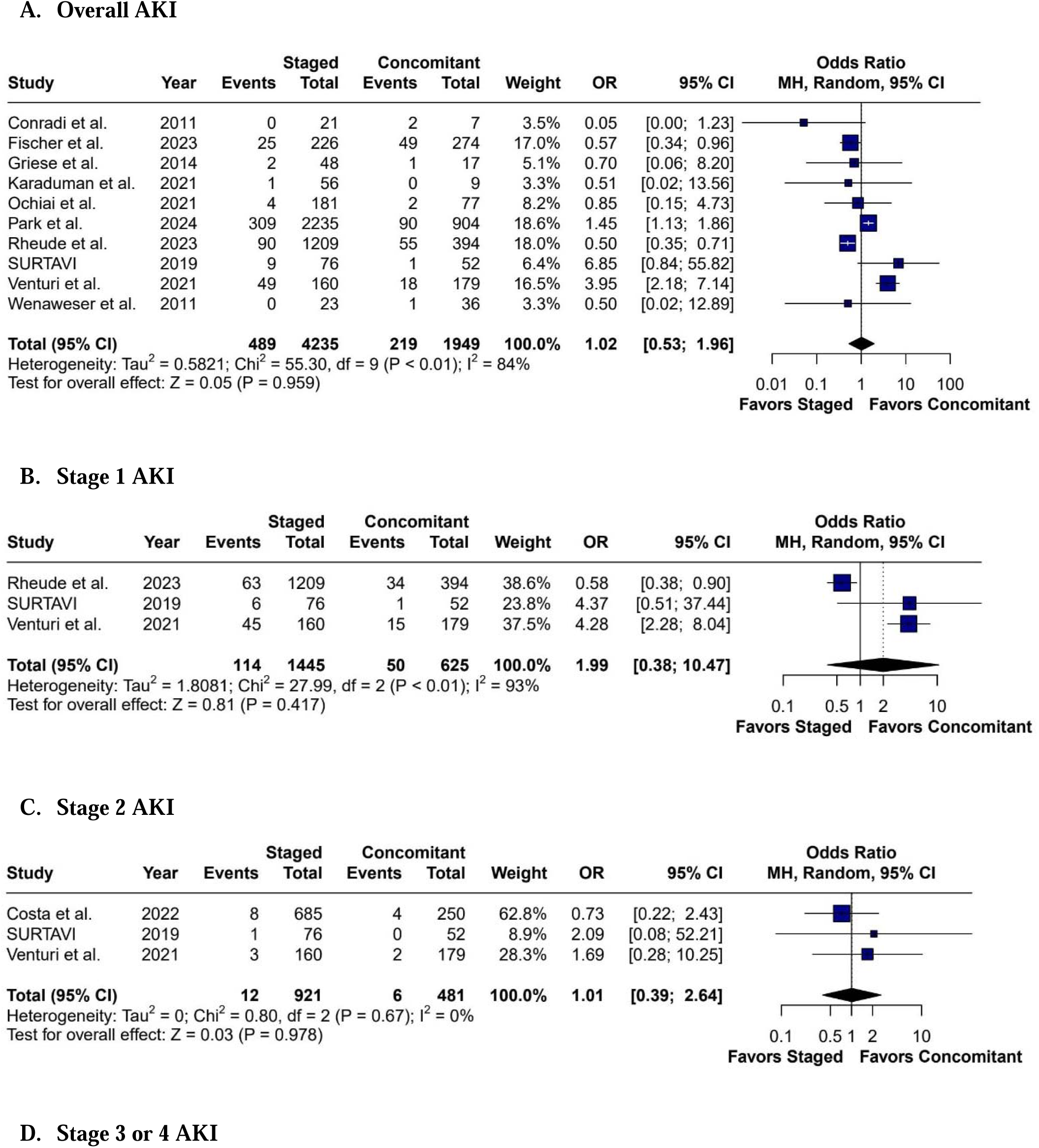

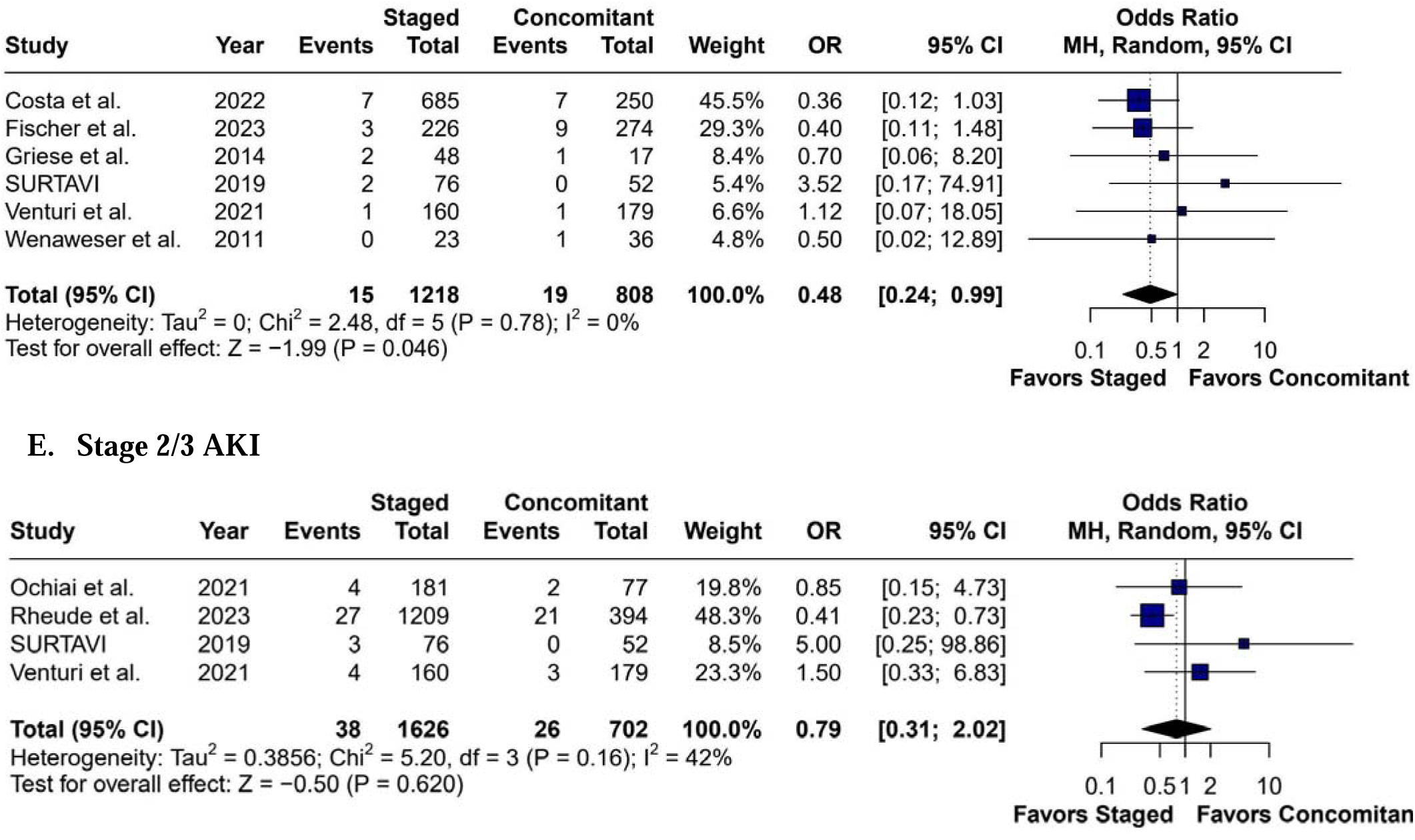

**Conclusion:** While staged PCI does not consistently reduce CI-AKI in patients undergoing TAVR, it may offer potential benefits for more severe kidney injury (stages 3/4). Given the observed heterogeneity, large-scale randomized controlled trials are essential to establish the relationship between procedural timing and renal outcomes.

## INTRODUCTION

The aging global population has experienced an increase in cardiovascular diseases, particularly aortic stenosis (AS) and coronary artery disease (CAD). These conditions share critical pathophysiological characteristics, including common risk factors such as hypertension and hypercholesterolemia. As they share similar pathophysiology involving pro-inflammatory states and lipoproteins, nearly half of all patients undergoing aortic valve intervention for AS have been shown to have concurrent CAD^1,2^. When untreated, the health and quality of life of patients with AS and CAD will be impacted as they eventually have impaired cardiac function, and eventually heart failure and death^3,4^. However, CAD has also been shown to be more indicative of comorbidity and surgical risk, rather than being an independent factor for poorer prognosis in those undergoing TAVR^2^.

Advances in minimally invasive surgical techniques have transformed treatment paradigms. Transcatheter aortic valve replacement/implantation (TAVR/TAVI) offers a less invasive alternative to traditional surgical aortic valve replacement (SAVR)^5^. At the same time, percutaneous coronary intervention (PCI) provides a comparable approach to coronary artery bypass grafting (CABG) for managing coronary artery disease. Despite these technological innovations, the optimal sequencing of TAVR and PCI remains a critical clinical challenge.

The complexity of managing patients with both AS and CAD extends beyond procedural considerations. Untreated conditions can lead to progressive cardiac dysfunction, increased morbidity, and potentially fatal outcomes. Current clinical guidelines typically recommend PCI for coronary stenosis exceeding 70% in proximal segments, yet the timing of interventions continues to be a subject of significant debate ^4,6^.

Existing literature presents conflicting perspectives on the optimal approach^2^. Some studies suggest that performing PCI first may risk decompensation due to untreated aortic stenosis. In contrast, others argue that prioritizing TAVR might increase myocardial infarction risk due to unaddressed coronary disease^9^. Critically, most existing research comprises observational studies with limited sample sizes and heterogeneous methodologies^2^.

Renal outcomes represent a particularly crucial consideration in this context. Renal impairment is both common in patients with heart failure and leads to increased mortality risk {Smith, L. Grace, 2006}. Both TAVR and PCI involve contrast media administration, which can precipitate contrast-induced acute kidney injury (CI-AKI)^2,5^. This risk is particularly pronounced in the typical patient population—elderly individuals and high-risk candidates with pre-existing renal vulnerabilities ^10^. The potential for contrast-induced nephropathy adds another layer of complexity to procedural decision-making.

Previous meta-analyses have yielded inconsistent results. While some studies demonstrate significant differences in renal outcomes between staged and concomitant procedures, others find no statistically significant variations^5,8^. This inconsistency underscores the need for comprehensive, systematic investigation.

Our study aims to address these critical gaps by systematically comparing the renal outcomes of staged versus concomitant TAVR-PCI procedures. Through a comprehensive meta-analysis, we seek to provide nuanced insights into the impact of procedural timing on acute kidney injury across different severity stages.

## METHODS

This meta-analysis was conducted and reported in accordance with the Cochrane Collaboration Handbook for Systematic Reviews of Interventions and the Preferred Reporting Items for Systematic Reviews and Meta-analyses (PRISMA) reporting guideline.^19,20^ The study protocol was prospectively registered in the International Prospective Register of Systematic Reviews (PROSPERO) database (CRD420261394979).

### Study Eligibility

Studies were eligible if they (1) were peer-reviewed full-text publications, (2) included adult patients (>18 years) with severe AS and CAD undergoing both PCI and TAVR; and (3) reported post-procedural renal outcomes. Studies were excluded if outcomes were not reported separately for the staged (SS) and concomitant (CC) PCI-TAVRstrategies.

### Search Strategy and Data Extraction

We systematically searched PubMed, Embase, and Cochrane Librairy on May 5, 2026. The primary outcome was the incidence of AKI according to itsdifferent stages. The complete search strategy and endpoint definitions are provided in Supplementary Tables S1 and S3, respectively. Study selection and data extraction were performed independently by at least two reviewers, with disagreements resolved by consensus.

### Quality Assessment

The methodological quality of the included observational studies was assessed using the Cochrane Risk of Bias in Non-randomized Studies of Interventions (ROBINS-I) tool.^21^ Publication bias was evaluated by visual inspection of funnel plots and Egger’s regression test.

### Statistical Analysis

Risk ratios (RR) with 95% confidence intervals (Cis) were calculatedusing the inverse- variance method. Between-study heterogeneity was assessed using I2 statistics, Cochran’s Q test, and Tau^2^ estimated with the restricted maximum-likelihood method. Heterogeneity was categorized as low (I^2^ = 0–25%), moderate (26–50%), or high (>50%). Leave-one-out sensitivity analyses were performed to evaluate the robustness of the findings and account for variability in study definitions. Meta-regression analyses explored the association between the incidence of AKI and the prevalence of chronic kidney disease (CKD). All analyses were performed using random-effects models in R version 4.2.1 (R Foundation for Statistical Computing).

## RESULTS

Our systematic review and meta-analysis included eleven retrospective observational studies comprising 7,119 patients with SAS who underwent either staged or concomitant PCI and TAVR procedures^3,9,12–20^. Comprehensive patient baseline characteristics and study/procedural details are provided in **Tables 1 and 2**, respectively.

**Table 1:**
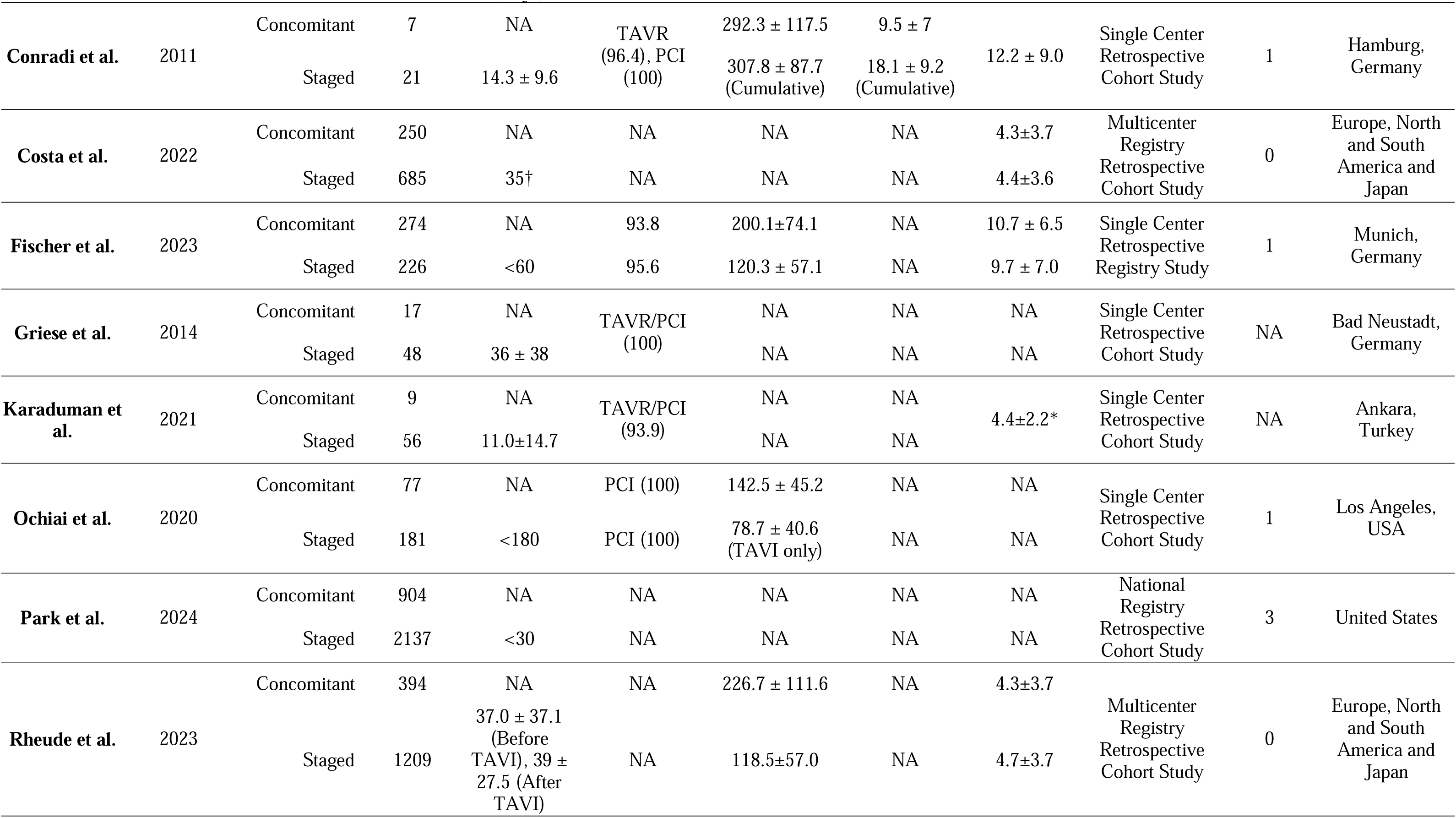
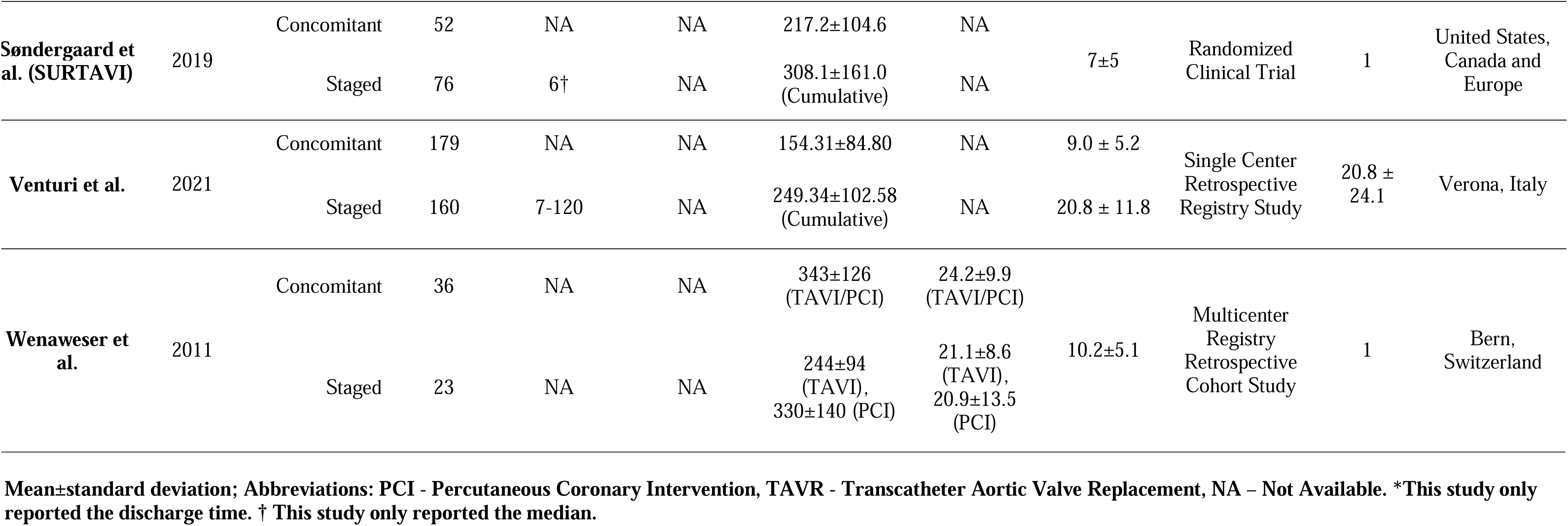
Study/Procedural Characteristics, Ordered by Primary Author.

**Table 2.**
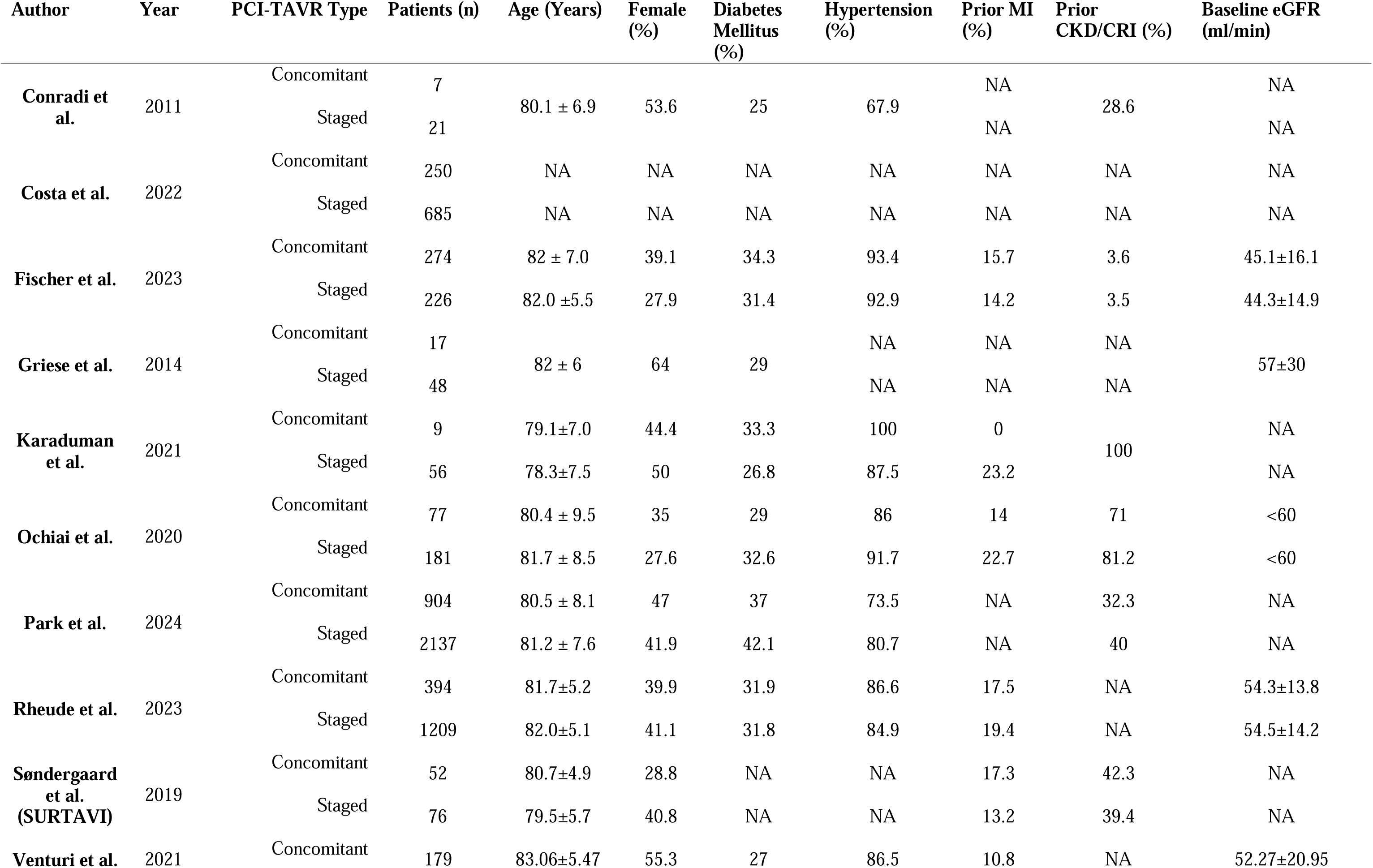
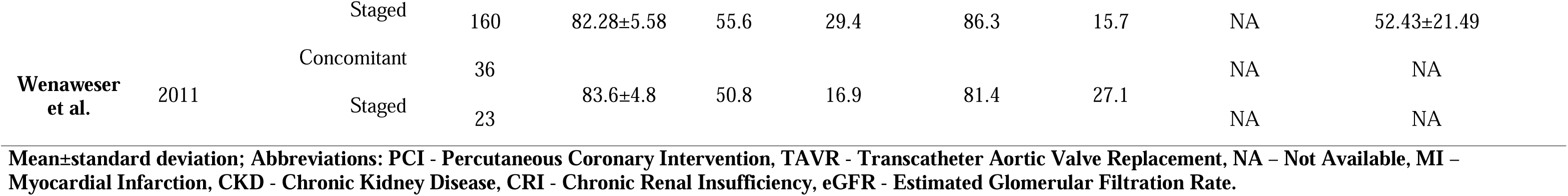
Patient Baseline Characteristics, Ordered by Primary Author.

The primary analysis revealed no significant reduction in overall CI-AKI incidence when comparing staged versus concomitant PCI (OR 1.02; 95% CI 0.53 to 1.98; p = 0.959; **Figure 1A**). Subgroup analysis revealed no significant differences in stage 1 (OR 1.99; 95% CI 0.38 to 10.47; p = 0.417; **Figure 1B**) or stage 2 CI-AKI (OR 1.01; 95% CI 0.39 to 2.64; p = 0.978; **Figure 1C**). However, a statistically significant difference emerged for stage 3/4 CI-AKI, favoring the staged approach (OR 0.48; 95% CI 0.24 to 0.99; p = 0.046; **Figure 1E**).

### Quality Assessment

The Risk of Bias in Non-randomized Studies (ROBINS-I) tool was used for quality assessment, and a significant proportion of the studies were considered at moderate to serious risk of bias as detailed in **Supplementary Figure S3**. Funnel plot analysis revealed asymmetrical distribution for stages 1 and 2/3 AKI, as illustrated in **Supplementary Figure S1**. Egger’s test indicated potential publication bias for stage 2/3 (p = 0.0455) and stage 3/4 AKI (p = 0.0530).

### Sensitivity Analyses

Due to moderate to high heterogeneity across all outcomes, we conducted a leave-one-out sensitivity analysis to validate the robustness of our results, and it showed that omitting the Venturi et al. study resulted in an OR adjustment of 0.24 for the overall AKI endpoint. Detailed sensitivity analysis results are presented in **Supplementary Figure S2**. Meta-regression analysis did not reveal a significant relationship between volume of contrast media and incidence of acute kidney injuries in both procedures (**Supplementary Table S2**).

## DISCUSSION

The landscape of cardiovascular interventions continues to evolve, with TAVR and PCI emerging as critical treatments for patients with SAS and CAD^3,5,8,20,21^. Our systematic review suggests that staged PCI did not significantly reduce the overall incidence of CI-AKI across various stages, except for a statistically significant benefit observed in the most severe stage 3/4 AKI subgroups. These results indicate that while spacing out contrast usage does not universally mitigate renal risks, it may provide meaningful protection for patients most susceptible to severe renal impairment.

While the ongoing TAVI-PCI trial is comparing PCI before and after TAVI, there are no large-scale randomized controlled trials to systematically assess outcome differences between these two approaches to the timing of PCI and TAVR. Currently, the number of patients with CAD that require TAVR is highly variable, ranging between 15% and 81%^2,22^. With evidence demonstrating increased MI risks in patients with untreated CAD after valvular interventions, appropriate management of CAD in presence of valvular pathologies is critical^4^. However, despite CAD in TAVR conferring poorer outcomes, PCI before TAVR did not demonstrate significant improvements^1,7,23^. Given the lack of high-quality evidence, the latest American College of Cardiology and American Heart Association outlined that the timing of CAD interventions in patients undergoing valve interventions is quite variable^4^. These would typically be driven by various clinical factors, including the presence of symptoms like angina, the ability and suitability for dual-antiplatelet therapy before TAVR, as well as the anatomical location and complexity of the vessel. The evidence, though non-randomized, was considered in the existing guidelines and was also noted to be safe and feasible. In particular, while it also noted that staged approach benefited from using lower contrast volumes to reduce renal failure risks, the timing of pre-TAVR PCI is still regarded as controversial^4,5^. In view of this, this systematic review aimed to evaluate the available evidence to date to better inform clinical practice until large-scale RCTs can be undertaken for more definitive conclusions.

In general, existing studies agree that PCI is both a safe and feasible approach to manage concomitant CAD in patients undergoing TAVR^5^. A network meta-analysis by Altibi et al., totaling ∼12,000 patients across twenty-two observational studies and two clinical trials, demonstrated no significant differences across 30 day all-cause mortality (OR: 1.19; 95% CI: [0.91–1.55], *P*= 0.20), 30 day cardiac mortality (OR 1.11; 95% CI: [0.55–2.26], *P*= 0.77) and 12 month mortality (OR 1.20; 95% CI: [0.59 – 2.44], *P*= 0.61)^24^. Of the two clinical trials, the first multicenter RCT (the ACTIVATION study) evaluating the impact of PCI on TAVI was completed in 2021 and agreed with existing observational literature on similar incidence of stroke, myocardial infarct, or AKI^25^. However, it too did not specifically evaluate the timing of PCI on renal outcomes. This evidence supports the overall efficacy but does not specifically address the question of optimal timing of PCI/TAVR. An important aspect of this is the independent effects of AKI on mortality. With renal function being vital to many physiological processes, especially during post-operative recovery, it is expected that the severity of renal impairment correlates with the increase in morbidity^26^. Hospitalized patients with AKI had an in-hospital mortality of 5.1% for stage 1 AKI, which increases to 13.7% for stage 2 and 24.8% for stage 3. Post-TAVR AKI was also observed to increase 3-year mortality, with a hazard ratio of 1.67 (95% CI 1.35-2.06)^13^.Therefore, this constitutes a vital consideration for all those undergoing cardiac procedures as many are already predisposed to increased morbidity and mortality.

Three of the studies in the same meta-analysis also reported on renal failure^5^. Overall, the renal risks were higher in the concomitant group than staged PCI (6.33; 95% CI [1.19, 33.64]; p=0.01). One of the studies, by Griese et al., demonstrated contrary evidence when comparing 16 staged and 17 concomitant interventions. All-cause mortality (p=0.01), cardiovascular mortality (p=0.01), and myocardial infarct (p=0.01) differed significantly between PCI with TAVR group, compared to TAVR alone^14^. However, when staged and synchronous PCI/TAVR cohorts were compared, none of the outcomes reached statistical significance (p>0.05). While AKIN 3 was specifically evaluated and demonstrated no significance either (p=1.00), only few developed in the staged (n=2; 4%) and concomitant groups (n=1; 6%). As a result, one of the key recommendations were that staged PCI is a better approach for those with CKD, at increased risks of renal failure, or for those with complex lesions whose PCI would likely take longer^5^. This was supported by Fisher et al., which compared 274 concomitant approaches against 226 staged and demonstrated increased odds of developing AKI (OR 1.74; 95% CI 1.03-3.00; p=0.041)^13^. Other smaller studies also supported staged approach, such as the study by Conradi et al., where 21 patients underwent concomitant PCI and 7 underwent staged intervention^3^. While only two developed acute renal failure in the concomitant group, one had pre-existing renal insufficiency. This supports the notion that the staged approach may be preferable in patients with impaired renal function. It should be noted that the OR for these smaller studies only included 60 concomitant and 92 staged cases and demonstrated non-significant differences in their respective studies, with ORs ranging between 1.99-19.55^3,14,20^. Therefore, given the low incidence of renal failure in these studies, impactful change on clinical practice will require more substantial evidence like large-scale RCTs.

While many studies have only reported renal failure or stage 3 equivalent AKI, the dose of contrast agent should also be evaluated as less severe cases of renal impairments may not have been captured due to the classification method. The theoretical contrast agent dose was expected to be much higher in concomitant procedures (mean difference 83.6 mL; 95% CI [50.47; 116.73]; p<0.00001) as the fluoroscopy time is significantly longer (18.1±9.2 vs 9.5±7.0 min; p=0.03)^3,5^. Therefore, the current consensus is that this be most relevant to those with renal impairments as the split dosage would reduce renal sequelae. This review of eleven studies showed that staged PCI does not significantly reduce the overall CI-AKI risks (OR 1.02; 95% CI 0.53-1.98; p=0.959). Furthermore, the mitigation of the larger doses used in concomitant procedures did not appear to affect the likelihood of developing stage 1 (OR 1.99; 95% CI 0.38, 10.47); p=0.70) or 2 AKI (OR 1.01; 95% CI 0.39, 2.64; p=0.70). Notably, only the stage 3/4 CI-

AKI risks reached statistical significance (OR 0.48; 95% CI 0.24-0.99; p=0.046). This potentially supports the notion that generally patients undergoing concomitant procedures do not result in renal impairment, whereas CI-AKI would develop in the more susceptible cohort, such as those with pre-existing renal impairment, and to a more severe degree. In the studies eligible for analysis, the 7 included significant proportion of patients with pre-existing renal impairments. These ranged from mild to significant, including some who were already under renal replacement therapy. Therefore, the recommendation based on our findings is that a staged approach provides considerable benefits over concomitant PCI/TAVR approach for patients with significant renal impairment prior to the procedure.

Another factor that contributed to the heterogeneity includes the differences in how AKI is defined. As reported in the RCT by Patterson et al., the baseline AKI incidence of patients undergoing TAVI are similar between TAVI alone (n=22; 19%) and pre-TAVI revascularization (n=17; 14.3%)^25^. These were reported in estimated glomerular filtration rates (eGFR). Among the various AKI definitions used by the analyzed studies, many also used the RIFLE criteria of risk, injury, failure, loss of kidney function, and end-stage renal disease while others used the acute kidney injury network (AKIN) stage 3^27^. The RIFLE criteria were an attempt to standardize the qualitative measurements and reporting of renal impairment as >200 different definitions of AKI can be used. RIFLE considers the serum creatinine or glomerular filtration rate changes in the severity of stages, from risk to ESRD. AKIN was developed by the Acute Kidney Injury Network group as a modified version of RIFLE, with Stage 1 being equivalent to RIFLE-Risk but with an increased serum creatinine of ≥ 0.3mg/dL (≥ 26.4 μmol/L), Stage 2 equivalent to RIFLE-Injury, and Stage 3 equivalent to RIFLE-Failure with the initiation of renal replacement therapy. Additional differences consider a different timeframe of creatinine change, with RIFLE being over 7 days and AKIN being <48 hours. While these may have influenced the classification of renal impairment in the evaluated studies, their application in critically ill patients have not been shown to differ significantly in sensitivity and accuracy of outcome prediction^28,29^. Within the reviewed studies, the pre-intervention renal function was reported in various formats. This included glomerular filtration rate, by CKD stage/ESRD, and whether patients were receiving dialysis. The continued non-standard reporting of renal function continues to make comparisons challenging. Similarly, most studies reported renal outcomes by AKI stage or RIFLE, which still left some ambiguity and was therefore addressed using leave-one-out sensitivity analysis. Nonetheless, while AKI stages 1 and 2 were not found to significantly differ with procedure, stage 3 AKI was of statistical significance. By using the leave-one-out analysis, the individual variation of each study was eliminated to identify the overall benefits of OR of 0.48. While this was statistically significant, there was also a wide confidence interval, ranging from 0.24 to 0.99. Therefore, although current literature suggests that these benefits support staged interventions, further investigation with larger more homogenous datasets and large-scale randomized controlled trials is still needed for quantify the benefits and impact in this patient cohort.

Additional factors to consider when interpreting these renal outcomes are the differences in patient cohorts’ characteristics. While all the included studies provided characteristics to some degree, the co-morbidities, especially diabetes, hypertension, previous myocardial infarcts, and other vascular disease, may affect outcomes significantly. These were reported to affect the majority of patients captured in the ACTIVATION study^25^. These comorbidities were not only prevalent among the patient cohort, but they also independently cause renal impairments and therefore must be considered in the interpretation of observational studies. Additionally, studies differed in the order of procedures, with some patients undergoing PCI before TAVR, and vice versa. As this is a clinical decision, this may also introduce selection bias and other clinical variables that were not captured in current observational studies. The differences in the location and number of diseased vessels may also affect these clinical decisions ^12^. Therefore, as the variations in the severity, order, and timing of PCI intervention can also affect outcomes, and these variables should also be evaluated appropriately in future randomized controlled trials. Additional biomarkers can also be considered in the diagnosis and monitoring of AKIs, including cystatin C, neutrophil gelatinase-associated lipocalin, interleukin-18, and kidney injury molecule-1^27^. Nonetheless, dedicated future studies evaluating renal sequelae following cardiovascular interventions should use and report standardized metrics to facilitate the interpretation and comparison of evidence.

Over the past decade, concomitant PCI and TAVI procedures have grown in popularity. The overall similar, and potentially favorable outcomes will continue to see their increasing adoption. Given the absence of randomized controlled trials on this, our systematic review and meta-analysis provides the most applicable evidence to support the management of CAD in patients undergoing TAVI. With many of these patients also having concurrent CKD, affecting up to 8% of the overall population, appropriate management of kidney function is an important consideration for clinicians as it is both more common in patients undergoing PCI/TAVR and is independently associated with higher mortality rates^30^. As such, given that many patients undergoing TAVI and PCI would have impaired baseline renal function, this review recommends the use of staged intervention for those with pre-existing renal impairment or at higher risks of AKI to reduce the risks. Additional perioperative management approaches are also necessary in this cohort, which includes optimizing contrast load through diuretics or ultrafiltration, use of drugs such as trimetazidine and using as little contrast dose as possible^31^. Specifically, the choice of contrast should also be considered, as iso-osmolar and low-osmolar contrast are preferable{Shroff, Gautam R., 2021; Lukwaro, Andrew, 2024}.

This was a rigorous systematic review and meta-analysis and included an extensive search for both relevant and previously reviewed studies. The search process was completed and reviewed independently by three authors, where a cardiologist clarified and mediated any discrepancies. Heterogeneity in study/procedural characteristics was moderate and was accounted for by conducting subgroup analyses while heterogeneity in patient characteristics was mild, specifically in those known to affect AKI outcomes (prior CKD, age). Random-effects model was also used to calculate confidence intervals for both inter- and intra-study variations. Strengths of the studies included exclusion of patients undergoing PCI for acute coronary syndrome (ACS) across the board improving heterogeneity.

The main limitation of this study is that there are few relevant studies, and where eligible, studies were limited by population size. Like other reviews with limited existing literature, publication bias cannot be eliminated. Additionally, as illustrated by existing guidelines, the clinical decision between staged or concomitant PCI/TAVR takes various factors into account and is not necessarily captured by these studies. Most notably, clinician preference, prior medical optimization, patients’ symptoms, and anatomical complexity of the lesions can significantly affect this choice and are rarely captured adequately in these observational studies. Inclusion criteria for kidney injury, staged PCI-TAVR and TAVR approach also varied considerably. Therefore, until large-scale, multi-center randomized controlled trials can be conducted to systematically compare these two approaches, the findings of this study should be applied on an individualized basis.

## CONCLUSION

The results of this meta-analysis did not find a difference in overall or stage 1/2 CI-AKI between staged and concomitant PCI-TAVR approaches with a slight benefit towards staged approach in stage 3/4 injury. A staged approach may be beneficial for patients with CAD and SAS with prior renal impairment. Further RCTs are needed to further investigate optimal timing strategies.

## Data availability statement

The data underlying this article are available in the article and in the Supplementary material online.

## Acknowledgements

Not applicable

## Author’s contributions

P.C. conceived and designed the study. P.C. and V.C. independently assessed the studies for possible inclusion and collected the data. P.C., and V.C. analyzed the data. P.C., V.C. and V.B. produced the first draft of the manuscript. P.G. made general supervision and was responsible for data interpretation and writing the final version. All authors approved the final version of the manuscript.

## Ethical approval

Ethical approval is not applicable. This study retrieved and synthesized data from already published studies.

## ABBREVIATIONS

AS: Aortic Stenosis
CA: Coronary Angiography
CI-AKI/CA-AKI: Contrast-Induced/Associated Acute Kidney Injury
CABG: Coronary Artery Bypass Grafting
CAD: Coronary Artery Disease
CKD/CKI: Chronic Kidney Disease/Insufficiency
CS: Concomitant Strategy
CI: Confidence Interval
ESKD/ESRD: End-stage Kidney/Renal Disease
LM: Left Main
MI: Myocardial Infarction
PCI: Percutaneous Coronary Intervention
PRISMA: Preferred Reporting Items for Systematic Reviews and Meta- Analysis
RCT: Randomized controlled trial
ROBINS-I: Cochrane Tool for Assessing Risk of Bias in Non-randomized Studies of Interventions
SAVR: Surgical Aortic Valve Replacement
SS: Staged Strategy
TAVI/TAVR: Transcatheter Aortic Valve Implantation/Replacement

