## Supplementary Materials for "Renal Outcomes of Staged Versus Concomitant Percutaneous Coronary Intervention and Transcatheter Aortic Valve Replacement: A Systematic Review and Meta-Analysis"

### **Supplementary Table S1: Definitions of Acute Kidney Injury across Studies**

| **Study** | **Year** | **Definition of CI-AKI** |
| --- | --- | --- |
| Conradi et al. | 2011 | NA - Acute Renal Failure requiring intensified medical treatment and temporary hemodialysis |
| Costa et al. | 2022 | VARC-2 Definition |
| Fischer et al. | 2023 | VARC-3 Definition in reference to KDIGO Definition |
| Griese et al. | 2014 | AKIN Classification (Modified RIFLE classification) |
| Karaduman et al. | 2021 | AKIN Definition |
| Ochiai et al. | 2020 | VARC-2 Definition |
| Park et al. | 2024 | ICD-10-CM Code N17 (Acute Kidney Injury) |
| Rheude et al. | 2023 | RIFLE Criteria |
| Søndergaard et al.  (SURTAVI) | 2019 | VARC-2 Definition |
| Venturi et al | 2021 | KDIGO Definition |
| Wenaweser et al. | 2011 | Modified RIFLE Classification |
| NA – Not Available, VARC – Valve Academic Research Consortium, AKIN – Acute Kidney Injury Network, RIFLE - Risk, Injury, Failure, Loss of kidney function, and End-stage kidney disease, KDIGO - Kidney Disease: Improving Global Outcomes | | |

### **Supplementary Table S2: Meta Regression Analysis – Acute Kidney Injury and Mean Cumulative Contrast Volume (mL)**

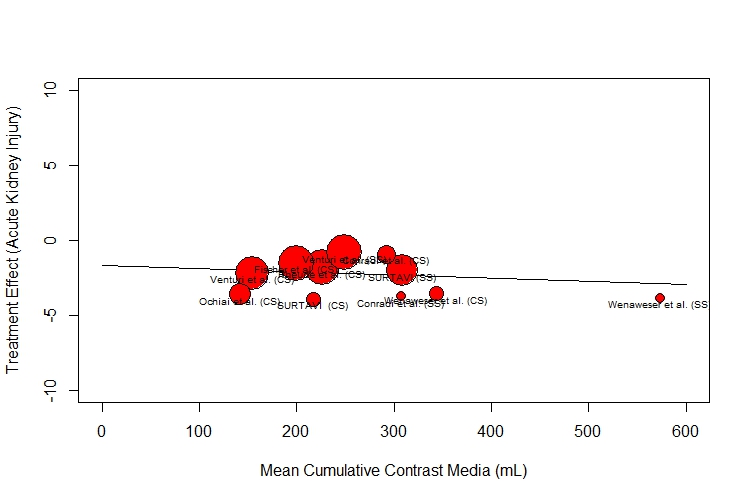

|  | Effect Estimate | p-value | I^2^ | Test for Residual Heterogeneity |
| --- | --- | --- | --- | --- |
| Intercept | -1.6801 | 0.0918 | 90.64% | p-val = < .0001 |
| Contrast Volume | -0.0021 | 0.5830 |  |  |
| CS – Concomitant PCI-TAVR, SS – Staged PCI-TAVR | | | | |

**Supplementary Table S3: Search Strategies**

| **Database** | **Search strategy** |
| --- | --- |
| **PubMed/**  **MEDLINE** | (“Coronary Artery Disease”[mh] OR CAD OR “Angioplasty, Balloon, Coronary”[mh] OR "percutaneous coronary intervention" OR PCI OR ((coronary) adj3 (revascular* OR interven* OR stent* OR angioplasty OR ballon))) AND  (“Transcatheter Aortic Valve Replacement” [mh] OR TAVR OR “Transcatheter Aortic Valve Implantation” OR TAVI OR ((transcatheter  OR trans-catheter OR “trans-cutaneous” OR trans-arterial OR transarterial OR transapical OR trans-apical) adj3 ("aortic valve") adj3 (implant* OR replace*))) NOT (letter[pt] OR comment*[pt]) NOT (animals[mh] NOT humans [mh]) |
| **EMBASE** | ('coronary artery disease'/exp OR 'coronary artery disease' OR 'percutaneous coronary intervention'/exp OR 'percutaneous coronary intervention' OR pci OR (coronary adj3 (revascular* OR interven* OR stent* OR 'angioplasty'/exp OR angioplasty OR ballon))) AND (tavr OR 'transcatheter aortic valve implantation'/exp OR 'transcatheter aortic valve implantation' OR 'tavi'/exp OR tavi OR ((transcatheter OR 'trans catheter' OR 'trans cutaneous' OR 'trans arterial' OR transarterial OR transapical OR 'trans-apical') adj3 ('aortic valve'/exp OR 'aortic valve') adj3 (implant* OR replace*))) NOT ([animals]/lim NOT [humans]/lim) NOT (letter:it OR editorial:it OR comment*:it OR ‘case report*’:it OR ‘case serie*’:it OR review:it OR ‘meta-analysis’:it OR 'conference abstract':it) |
| **COCHRANE** | (“coronary artery disease” OR CAD OR "percutaneous coronary intervention" OR PCI OR ((coronary) AND (revascular* OR interven* OR stent* OR angioplasty OR ballon))) AND (“Transcatheter Aortic Valve Replacement” OR TAVR OR “Transcatheter Aortic Valve Implantation” OR TAVI OR ((transcatheter  OR trans-catheter OR “trans-cutaneous” OR trans-arterial OR transarterial OR transapical OR trans-apical) AND ("aortic valve") AND (implant* OR replace*))) |

### **Supplementary Figure S1. Publication Bias Assessment**

A. **Overall AKI**

**Egger’s test (p-value = 0.7864371)**

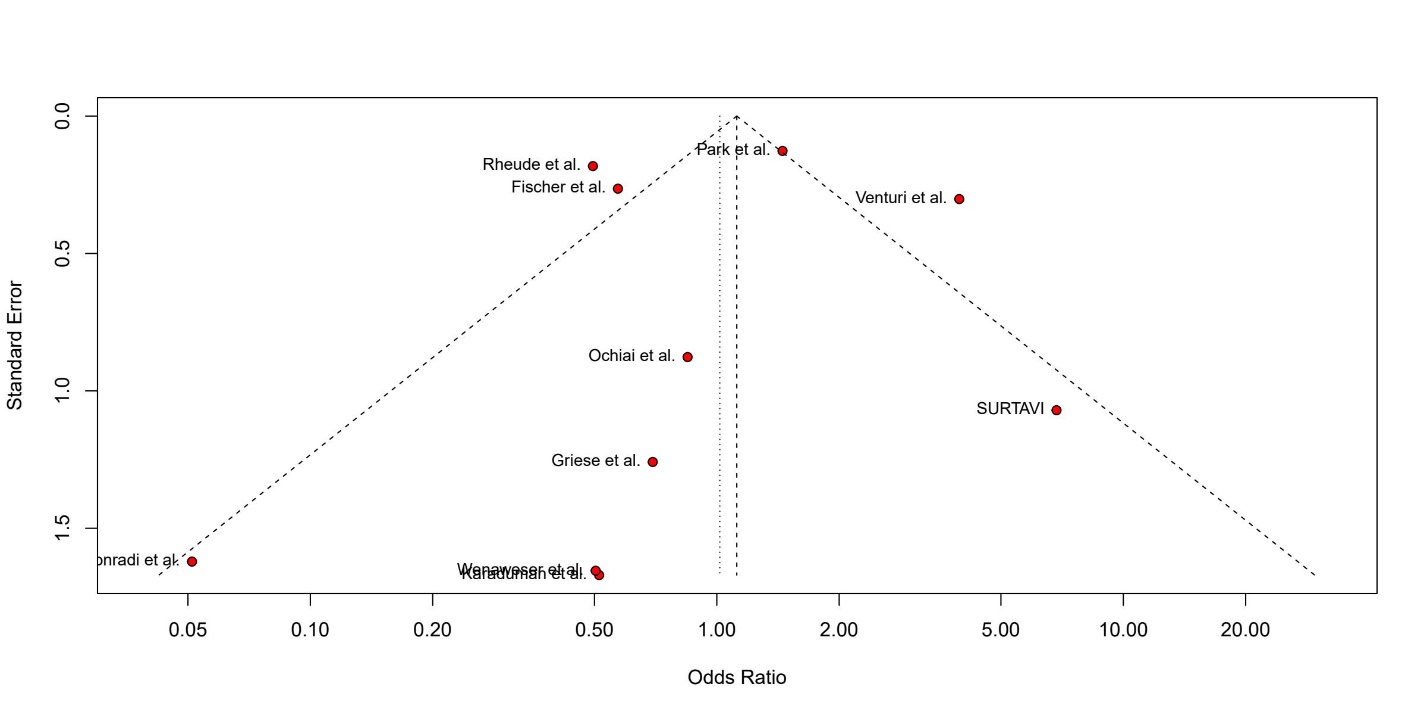

B. **Stage 1 AKI**

**Egger’s test (p-value = 0.6396095)**

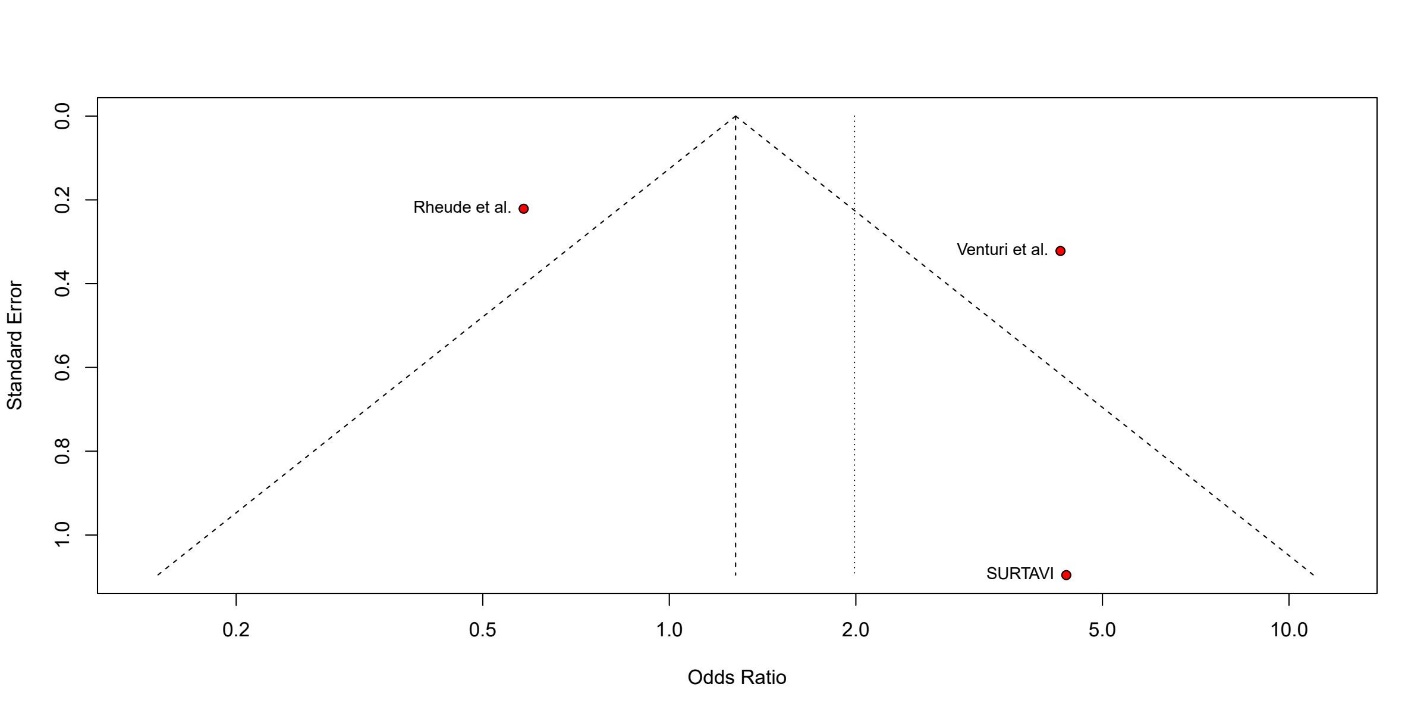

C. **Stage 2 AKI**

**Egger’s test (p-value = 0.3568132)**

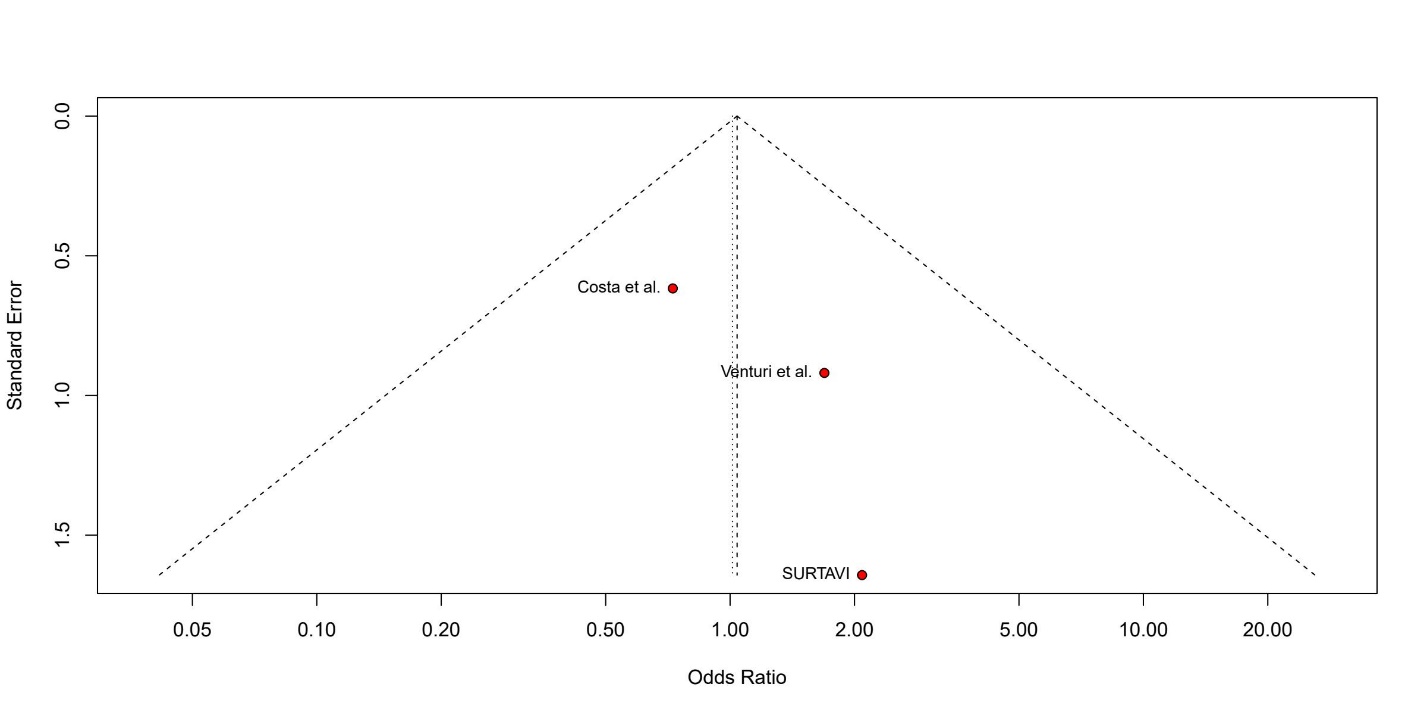

D. **Stage 3 or 4 AKI**

**Egger’s test (p-value = 0.05304349)**

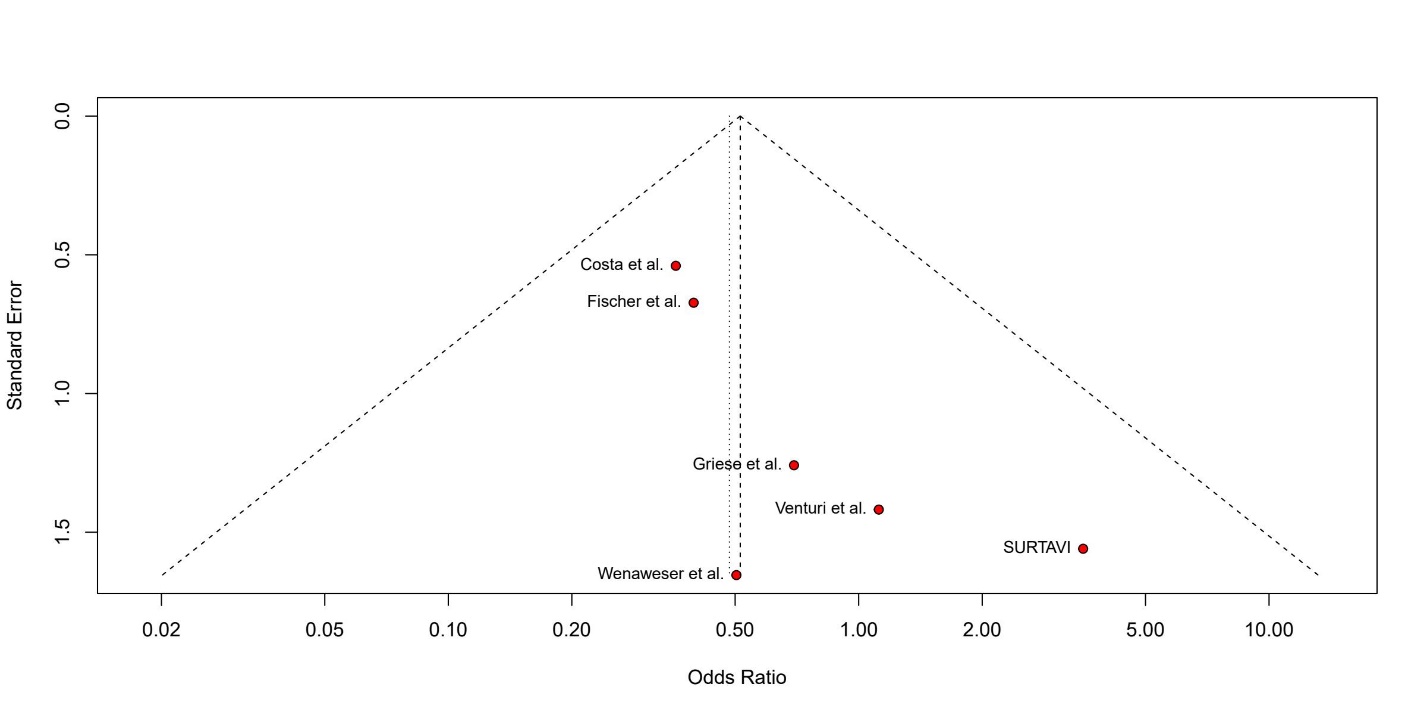

E. **Stage 2/3 AKI**

**Egger’s test (p-value = 0.04553117)**

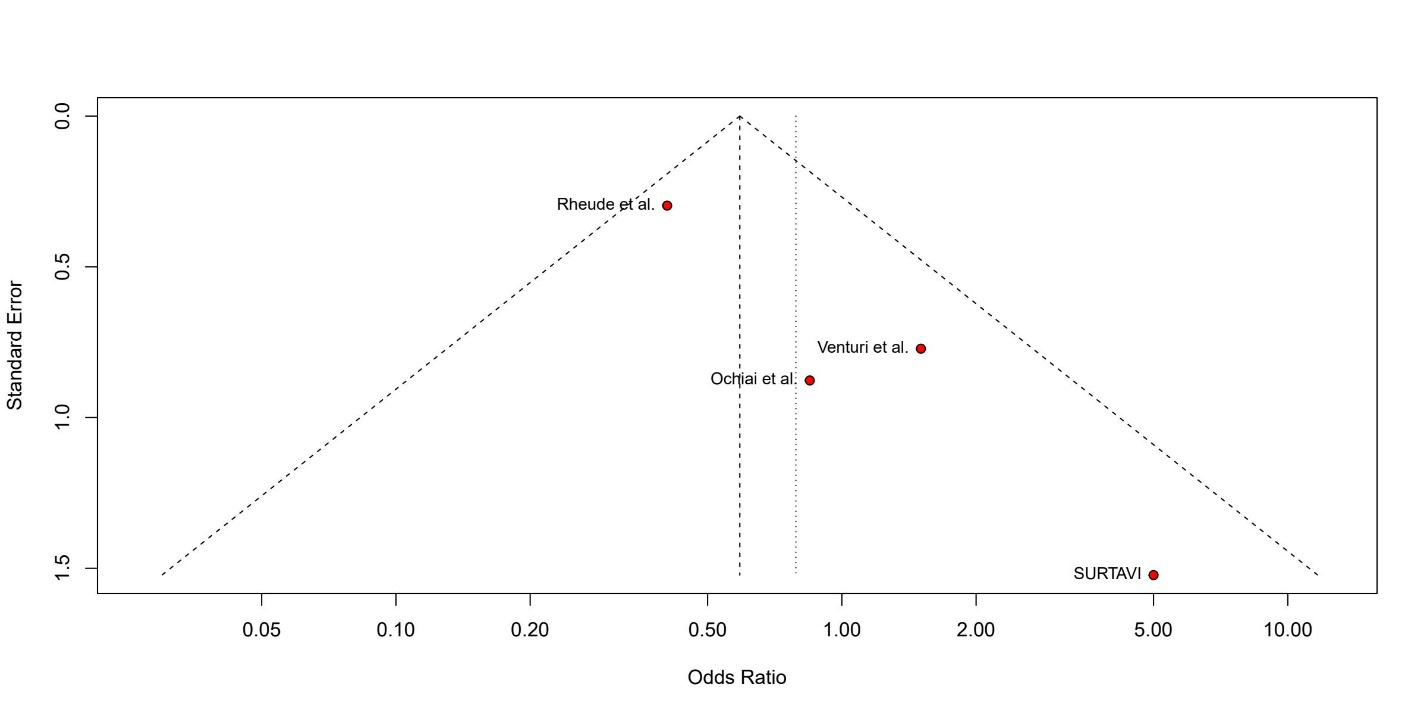

### **Supplementary Figure S2. Leave-one-out Sensitivity Analyses**

A. **Overall AKI**

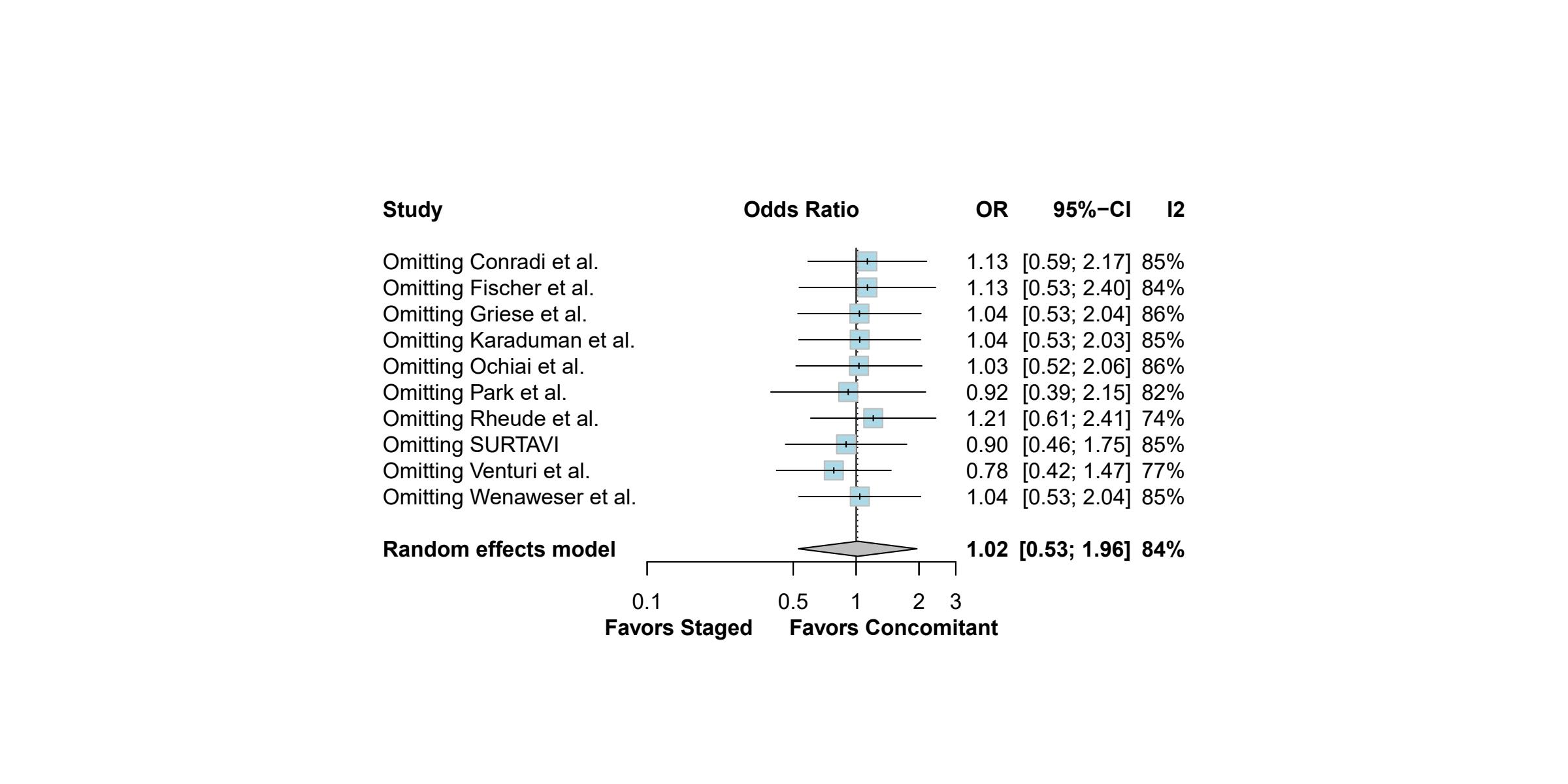

B. **Stage 1 AKI**

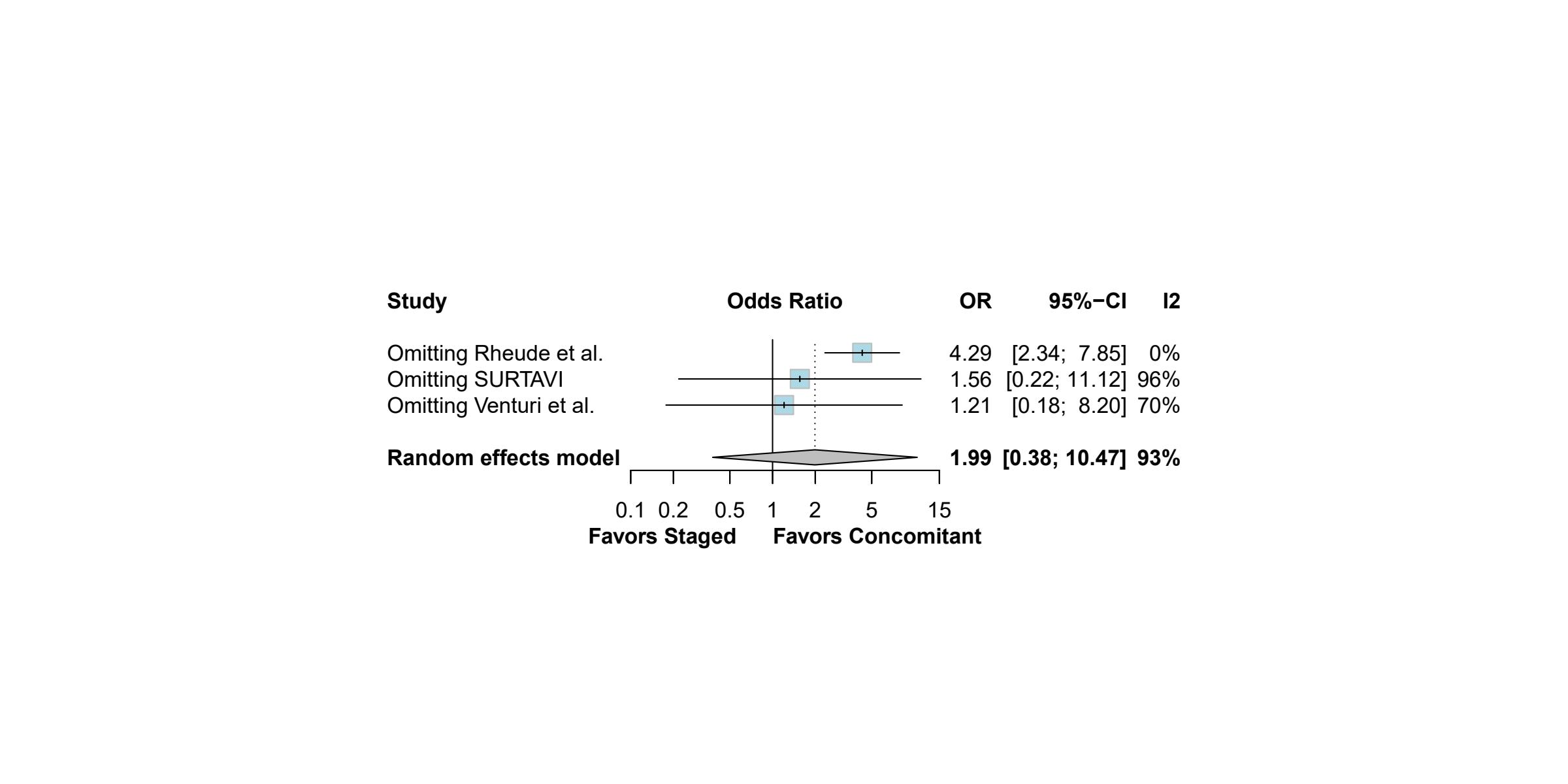

C. **Stage 2 AKI**

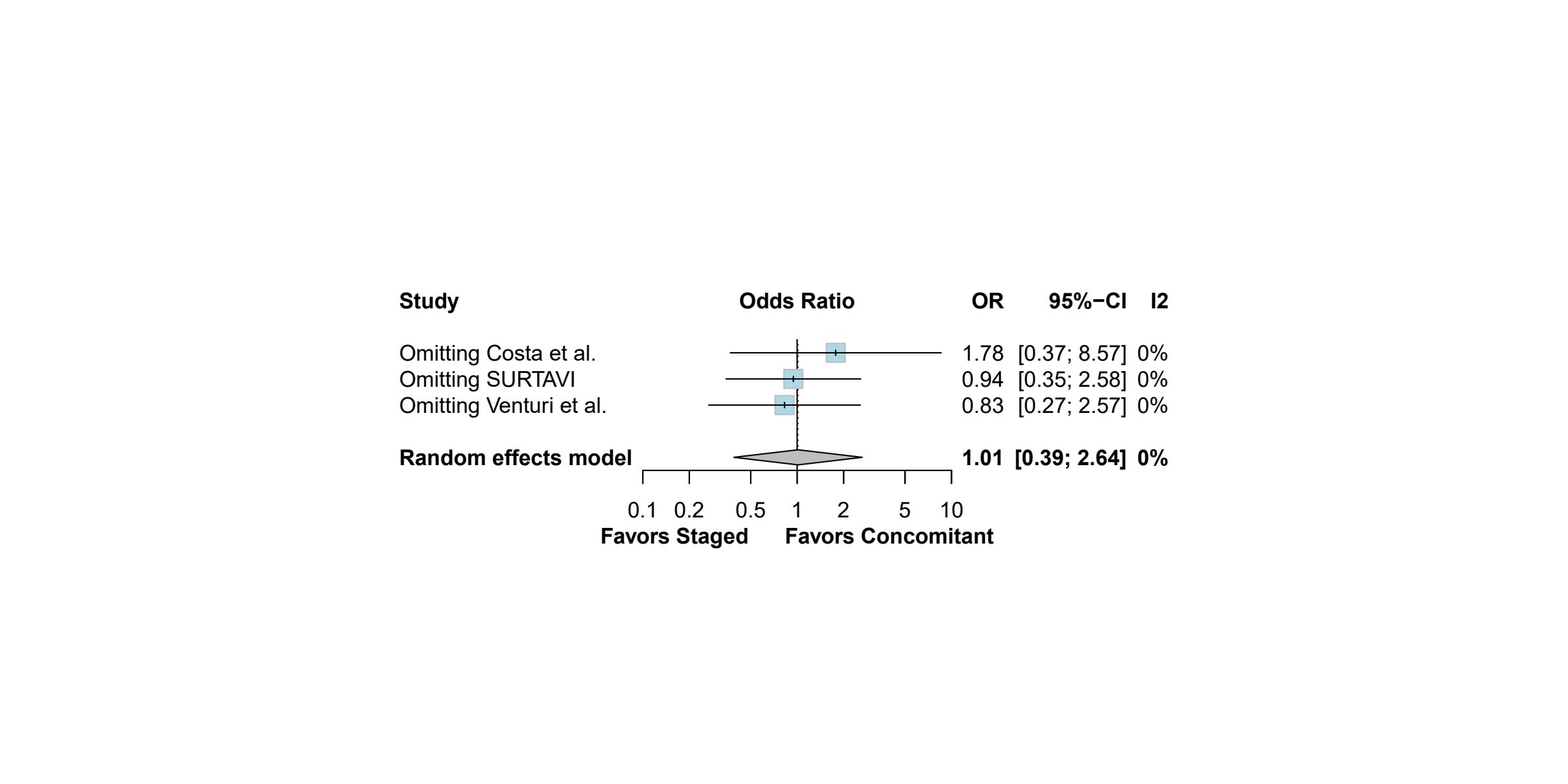

F. **Stage 3 or 4 AKI**

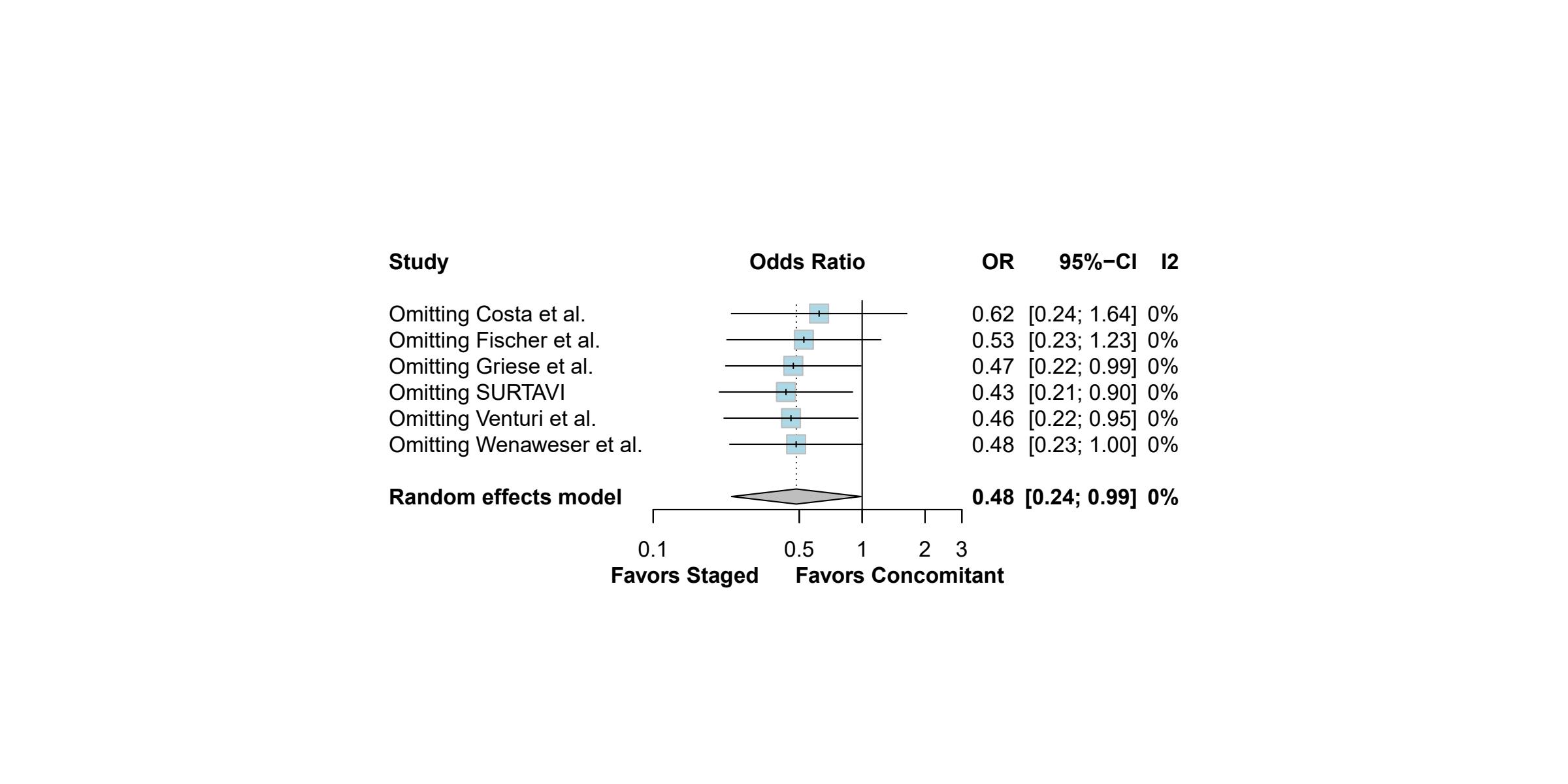

D. **Stage 2/3 AKI**

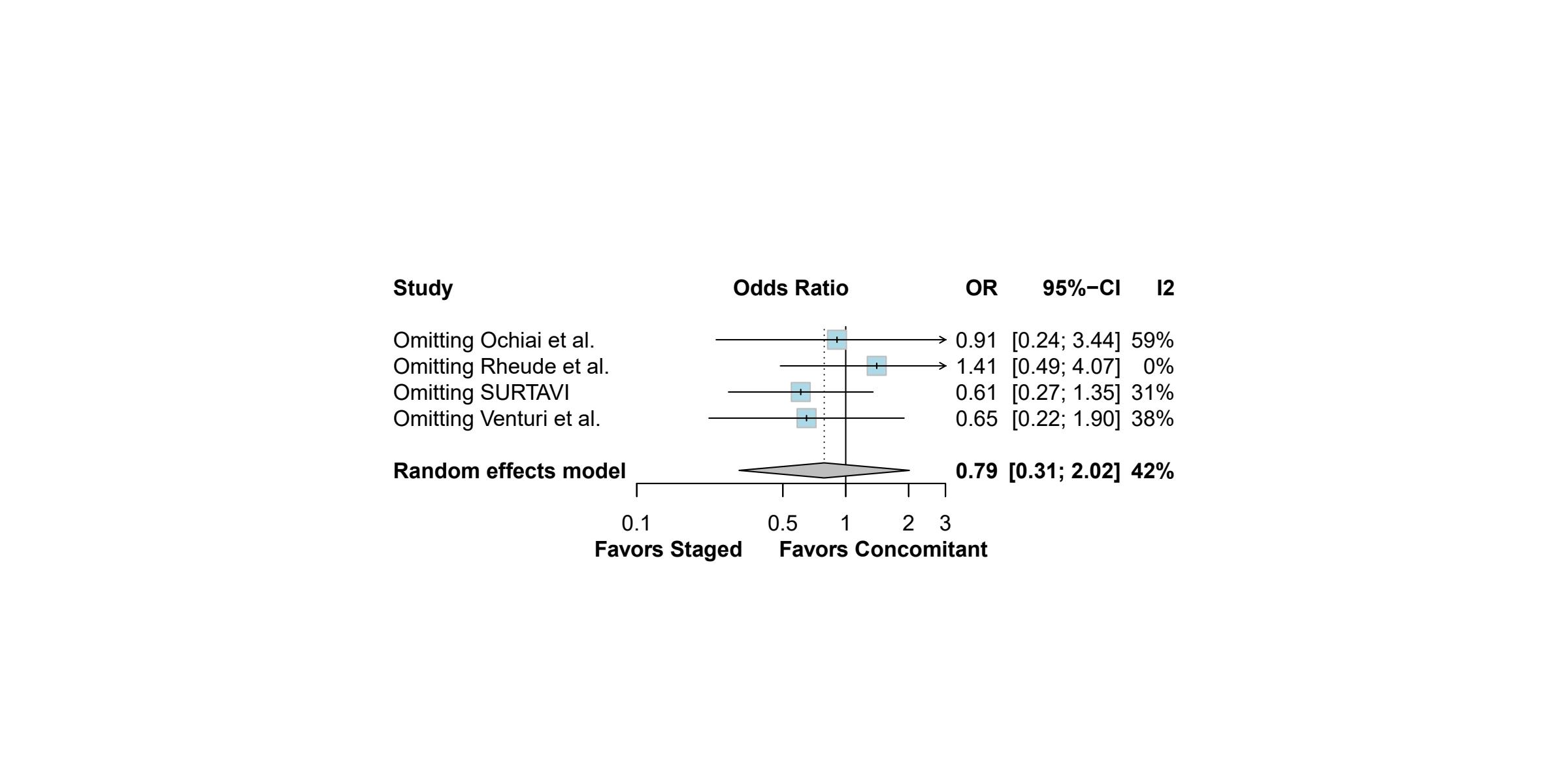

### **Supplementary Figure S3. Risk of Bias Assessment - Cochrane Tool for Assessing Risk of Bias in Non-Randomized Trials (ROBINS-I)**

A. **Summary**

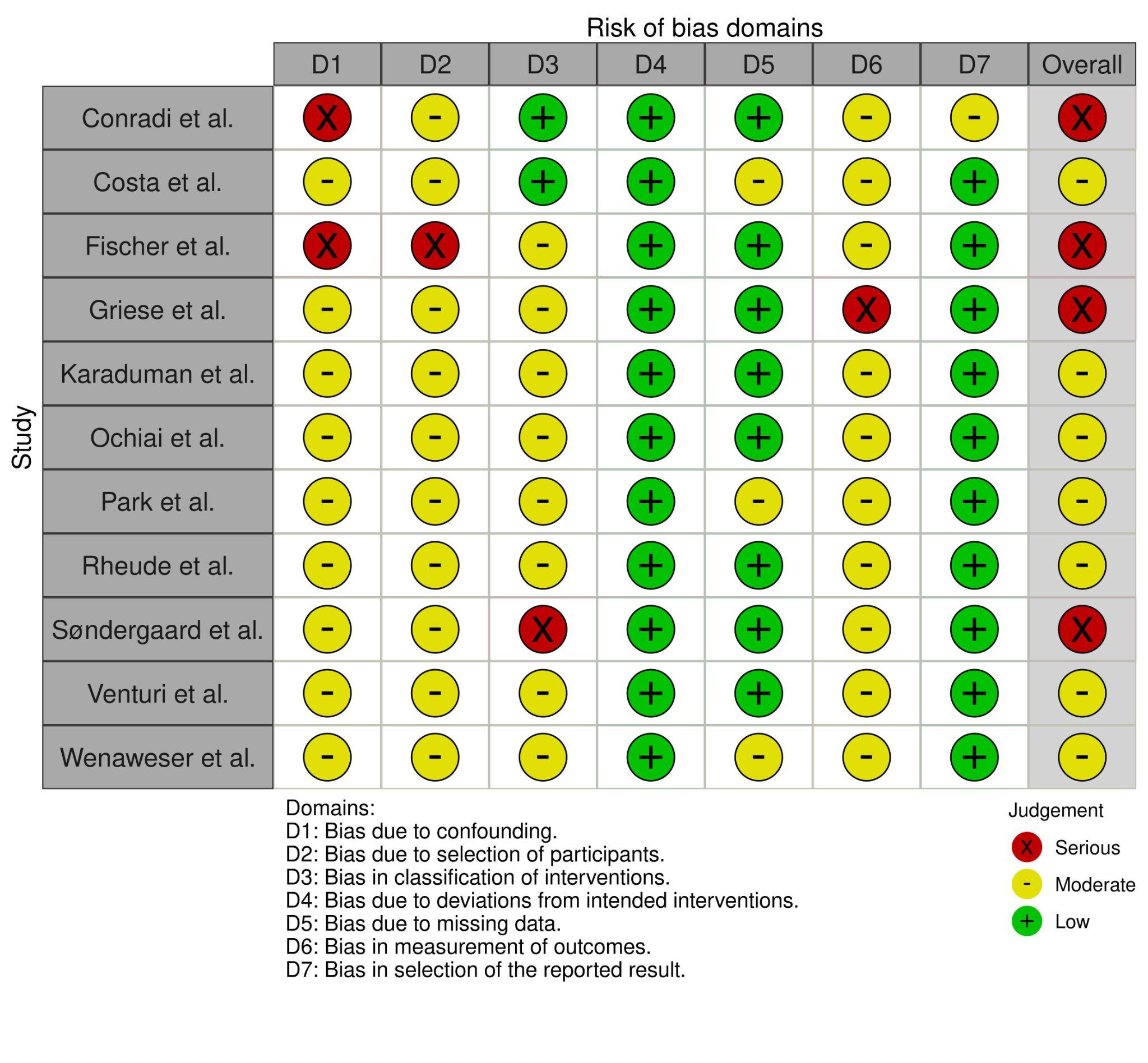

B. **Graph**

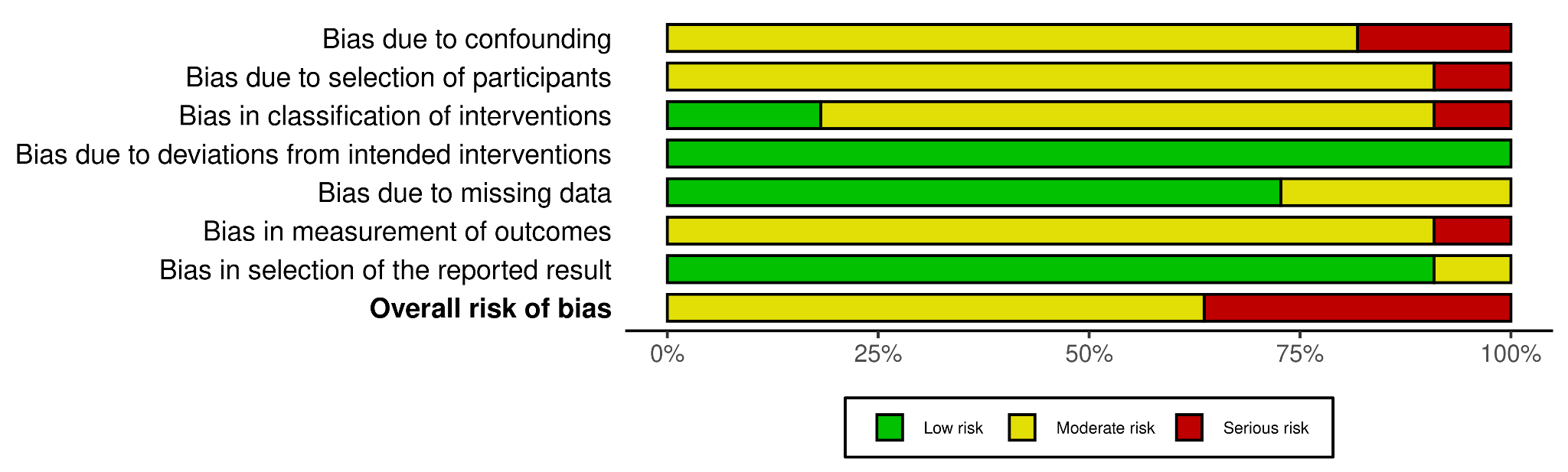
